# Cross-frequency phase-amplitude coupling in repetitive movements in patients with Parkinson’s disease

**DOI:** 10.1101/2021.07.26.21261085

**Authors:** Ruxue Gong, Christoph Mühlberg, Mirko Wegscheider, Christopher Fricke, Jost-Julian Rumpf, Thomas R. Knösche, Joseph Classen

**Affiliations:** Department of Neurology, Leipzig University Medical Center, Leipzig, Germany; Method and Development Group Brain Networks, Max Planck Institute for Human Cognitive and Brain Sciences, Leipzig, Germany

**Keywords:** Phase-amplitude coupling, Parkinson’s disease, bradykinesia, repetitive movement, motor control

## Abstract

Bradykinesia is a cardinal motor symptom in Parkinson’s disease whose pathophysiology is incompletely understood. When signals are recorded from the cortex or scalp at rest, affected patients display enhanced phase-amplitude coupling between β (13-30Hz) and broadband γ (50-150Hz) oscillatory activities. However, it remains unclear whether and how abnormal phase-amplitude coupling is involved in slowing Parkinsonian movements during their execution. To address these questions, we analyzed high-density EEG signals recorded simultaneously with various motor activities and at rest in 19 patients with Parkinson’s disease and 20 healthy controls. The motor tasks consisted of repetitive index finger pressing, and slow and fast tapping movements. Individual EEG source signals were computed for the premotor cortex, primary motor cortex, primary somatosensory cortex, and primary somatosensory complex. For the resting condition and the pressing task, phase-amplitude coupling averaged over the 4 motor regions and the entire movement period was larger in patients than in controls. In contrast, in all tapping tasks, state-related phase-amplitude coupling was similar between patients and controls. These findings were not aligned with motor performance and EMG data, which showed abnormalities in patients for tapping but not for pressing, suggesting that the strength of β-broadband γ phase-amplitude coupling during the movement period does not directly relate to Parkinsonian bradykinesia. Subsequently, we examined the dynamics of oscillatory EEG signals during motor transitions. When healthy controls performed the pressing task, dynamic phase-amplitude coupling increased shortly before pressing onset and decreased subsequently. A strikingly similar motif of coupling rise and decay was observed around the offset of pressing and around the onset of slow tapping, suggesting that such transient phase-amplitude coupling changes may be linked to transitions between different movement states – akin to preparatory states in dynamical systems theory of motor control. In patients, the modulation of phase-amplitude coupling was similar in (normally executed) pressing, but flattened in slow (abnormally executed) tapping compared to the controls. These deviations in phase-amplitude coupling around motor action transients may indicate dysfunctional evolution of neuronal population dynamics from the preparatory state to movement generation in Parkinson’s disease. These findings may indicate that cross-frequency coupling is involved in the pathophysiology of bradykinesia in Parkinson’s disease through its abnormal dynamic modulation.

## Introduction

Cross-frequency coupling between the phase of β oscillations and the amplitude of γ oscillations is widely considered as a pathophysiological biomarker for Parkinson’s disease.^1^ Recordings via subdural electrocorticography (ECoG), local field potentials (LFP), and even non-invasive EEG revealed enhanced β-γ phase-amplitude coupling (PAC) during the resting state of patients with Parkinson’s disease in both cortex^1-3^ and subthalamic nucleus (STN)^4^, as compared with either healthy controls or patients with non-movement disorders. Exaggerated β-γ PAC at rest in patients with Parkinson’s disease has been associated with motor impairment^1, 3-5^ and can be attenuated by deep brain stimulation of the STN or by dopamine replacement therapy.^6, 7^ Despite this correlative evidence, there is a possibility that enhanced β-γ PAC at rest is merely an epiphenomenon, or a surrogate marker of a distant pathophysiological mechanism and, therefore, not causally related to impaired motor performance. Studies on the role of PAC magnitude during movement in patients with Parkinson’s disease have not settled this issue. Movement-related PAC derived from LFP recordings from the STN and globus pallidus externus, as well as from ECoG recordings from sensorimotor areas in patients with Parkinson’s disease was reduced compared to resting state.^6, 8-10^ Furthermore, DBS-induced acceleration of movement during a complex reaching and tracking task was associated with reduced cortical movement-related PAC.^6^ These observations appeared to provide circumstantial evidence that bradykinesia of patients with Parkinson’s disease is related to persistently enhanced PAC during movement. However, PAC derived from scalp EEG during a verbally cued intermittent hand opening/closing task did not differ between patients in the off-medication state and healthy subjects.^5^ Even in the studies reporting persistent PAC enhancement during movement, the relationship between the strength of PAC and the motor impairment remained unclear since none of the studies reported whether the execution of the movements was actually impaired in patients with Parkinson’s disease.^1, 11^ Because abnormalities of movement-related PAC may not become apparent until kinematic abnormalities occur, the lack of information on kinematics means that it is unknown how PAC is involved in the pathophysiology of movement disorders.

Bradykinesia (slowness of motor execution) and movement amplitude decrement, evident when movements are performed repetitively, represent cardinal manifestations of the motor impairment of patients with Parkinson’s disease.^12^ Other movement parameters, such as grip force^13^ or reaction time^10^ do not reliably distinguish between patients and controls. Likewise, the tapping rate in self-paced tapping at the patients’ own pace^14^ or tapping paced by external sensory inputs^15, 16^ may be normal in patients with Parkinson’s disease. The co-occurrence of pathologically altered and intact movement parameters in patients with Parkinson’s disease allows us to elucidate the functional role of PAC in the pathophysiology of movement impairment by comparing different movement types. A unifying pathophysiological mechanism should reflect the behavioral pattern: that is, it should be abnormal during abnormal motor behavior and normal during normal motor behavior.

In previous work^3^, we have investigated the characteristics of exaggerated β-γ PAC during rest. Here, we report our analyses of oscillatory EEG activity during different types of voluntary repetitive movement tasks. Some of these tasks were chosen to specifically elicit the cardinal motor signs as revealed by the clinical examination. The study of various forms of abnormal and normal movement behavior in Parkinson’s disease may shed new light on the functional role of PAC in movement dynamics and the relationship between PAC and Parkinsonian motor impairment.

## Methods

### Participants

Nineteen patients with Parkinson’s disease (6 females, mean age: 60.9±10.8y) and twenty age- and sex-matched healthy controls (8 females, mean age: 62.6±7.9y) were recruited and included in the analysis of this study. The characteristics for patients have been described in detail previously^3^ and are provided in Supplementary Table 1. There was no significant age difference between patients and controls (p=0.527). The experiment was carried out under a practically defined ‘off-medication’ state (at least 12 hours overnight withdrawal of Parkinsonian medication). Written informed consent was obtained from all patients and controls according to the protocol approved by the local Ethics Committee at the Medical Faculty of Leipzig University (Reference number: 147/18-ek).

**Table 1.** Relationship between the movement-related change of PAC and change of β power

| Task | P1-P2 // T1-T2 | P2-P3 // T2-T3 | P3-P4 // T3-T4 | P4-P5 // T4-T5 |
| --- | --- | --- | --- | --- |
| Pressing | $R=0.13$, $p=0.429$ | **$R=0.62$, $p=0.001$** | **$R=0.42$, $p=0.017$** | $R=0.23$, $p=0.205$ |
| Slow tapping | **$R=0.50$, $p=0.003$** | **$R=0.42$, $p=0.017$** | $R=-0.09$, $p=0.568$ | $R=0.18$, $p=0.362$ |

\*p values have been FDR corrected for each task, all significant values after correction are marked in bold

### Movement recording

Pressing and tapping tasks (see below) were recorded by a custom-made device consisting of a force transducer and two photoelectric beam sensors (Fig 1A). The digital signal from the force transducer served to determine the real-time mechanical onset and offset of the pressing task. During the pressing task, the onset and offset of pressing were defined as the moments when the force exceeded or fell below a threshold of 1.3 N, respectively. The maximum force that the transducer could detect was 4.4N.

**Figure 1.**
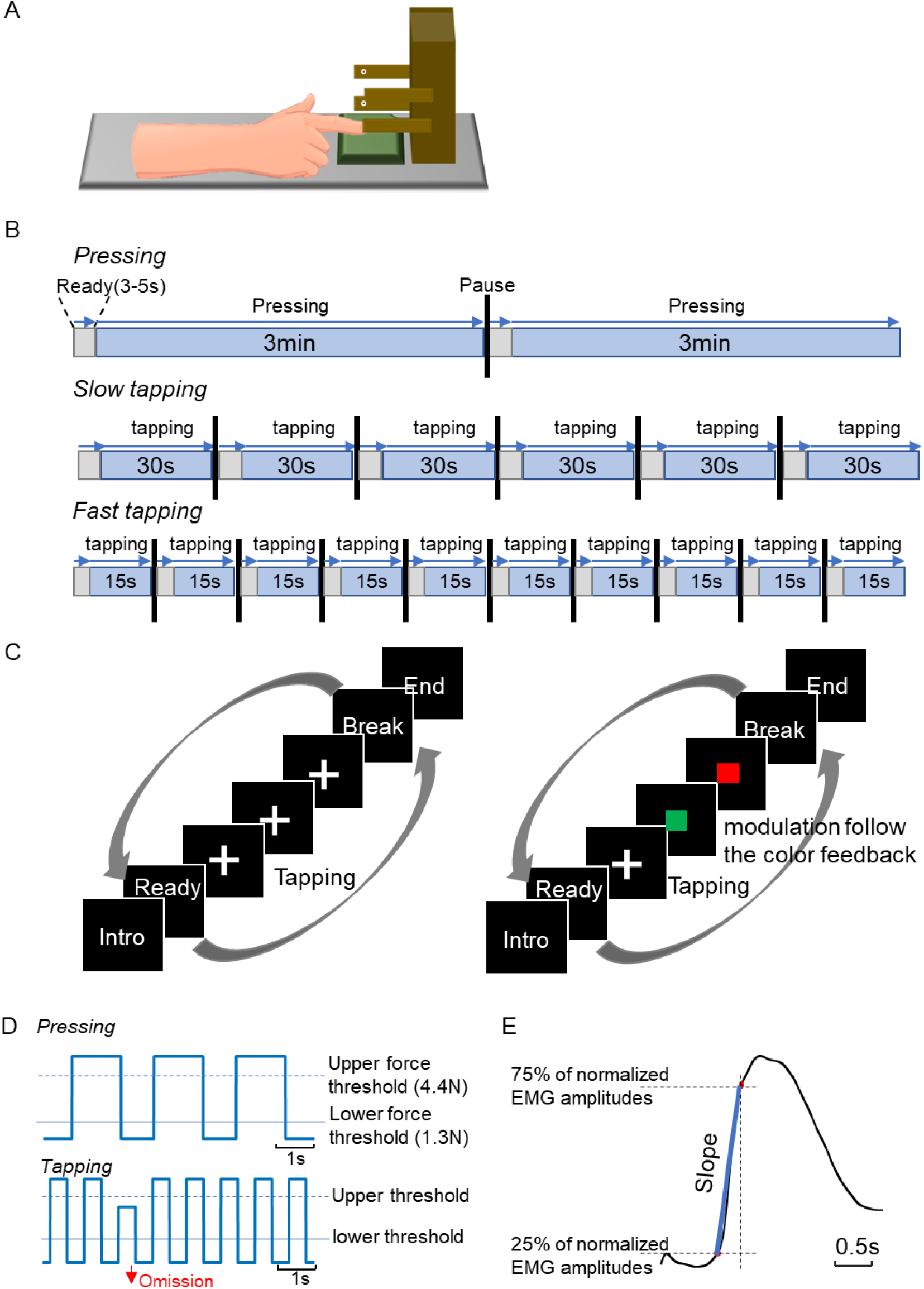
Experimental design. A. Schematic diagram of the experimental setup. The device for recording repetitive movements consisted of a transducer (green plate) recording pressing force and two photoelectric sensors (white) reporting the height of the extended index finger. B. Experimental protocol for the three movement tasks. Grey areas represent the short periods when subjects prepared to perform the task. Blue areas represent the active movement periods. C. Experimental design of the tapping tasks. There were two conditions for each tapping task. Left: Subjects performed repetitive tapping while looking at the fixation cross. Right: After each tapping cycle, subjects received color feedback indicating whether the extended index finger had reached the upper level. A green square indicated that the level of the preceding tapping movement had been at or above the upper photoelectric sensor. A red square indicated that the tapping height had been below the upper photoelectric sensor. Subjects were instructed to maximize green feedback by performing sufficiently extended tapping movements. D. Schematic representation of the force and amplitude trajectories during pressing (upper curve) and slow tapping (lower curve). In pressing, movement onset was defined as the moment when the force signal exceeded the lower force threshold (1.3N). No force signals were resolved above the upper threshold of 4.4N. During tapping, the online movement onset was defined as the moment when the index finger was extended above the light beam of the lower photoelectric sensor. An omission was recorded if the index finger was not extended high enough to reach or cross the light beam of the upper sensor. E. The estimation of EMG slope. It was defined as the slope of the line (blue) connecting the 25% and 75% percentile (red dots) of the normalized EMG signal closest to the mechanical onset of tapping.

The lower photoelectric sensor was placed at a height just above the finger when it was placed on the pressure sensing board, and the upper photoelectric sensor was placed at the height of the extended index finger in a position parallel to the table. During the tapping tasks, the real-time mechanical onset of index finger extension was defined as the time when the light beam of the lower photoelectric sensor was interrupted by the extended index finger. The upper photoelectric sensor was used to signal the elevation level of the extended index finger during tapping. In tapping movements, omissions were recorded if the index finger did not reach the upper photoelectric sensor level. Figure 1D illustrates how information of movement events was determined during the pressing and tapping tasks. The mechanical onsets recorded online were visually verified afterwards for mechanical or performance errors.

### Movement tasks

Participants were asked to perform three different voluntary movement tasks involving repetitive index finger actions: pressing, tapping at slow speed, and tapping at fast speed. We tested the performance of the patients on the hand side most severely affected by the disease, as indicated by the bradykinesia hemi-body scores (10 patients in the left and 9 patients in the right) in part III of the Movement Disorder Society Unified Parkinson’s Disease Rating Scale (MDS-UPDRS III). In healthy controls, the side for performing the movement tasks was pseudorandomly chosen to eventually match the respective sub-sample sizes of patients (10 controls in the left and 10 controls in the right). Before starting each trial, subjects were asked to place their arm on an armrest and place their index finger on the pressure sensing board. Participants were asked to start the index finger movements as soon as a white cross appeared at the center of the computer screen.

The experimental protocol is illustrated in Fig 1B. In the pressing task, subjects were asked to perform a self-initiated pressing and releasing movement using the index finger at a comfortable rate (2 trials of 3min each). In the slow tapping tasks (6 trials of 30s each, two blocks), subjects were instructed to perform tapping at a slow rate (‘tap at your own comfortable speed’). In the fast tapping tasks (10 trials of 12s or 15s each, two blocks [9 patients and 11 controls tapped for 12s per trial in the second block of fast tapping tasks]), subjects were instructed to perform tapping at their fastest speed (“tap as fast as possible”). Between trials and tasks, subjects were allowed sufficient time to rest, in order to minimize fatigue. During slow tapping and fast tapping tasks, subjects were asked to tap using their maximal index finger extension. Two different conditions (blocks) were designed for each slow and fast tapping task (Fig. 1C). The conditions differed in the absence or presence of feedback as to whether the extension of the index finger had met the upper-level criterion as detected by the upper photoelectric light beam. In the first condition (FB-), subjects were asked to perform repetitive tapping at the instructed tapping rate without receiving information about the tapping amplitude (Figure 1C, left). In the second condition (FB+), subjects were instructed to increase their index finger’s height according to the feedback if necessary. The feedback consisted of a colored square displayed on the screen after each tap whose color indicated whether the index finger extension had reached the upper threshold as reported by the upper photoelectric sensor (Figure 1C, right). Therefore, the motor tasks consisted of 5 conditions: repeated pressing at a slow rate, slow tapping without feedback, slow tapping with feedback, fast tapping without feedback, and fast tapping with feedback.

### Motor performance metrics

In each tapping task, motor performance was indexed by tapping rate, tapping variability, and the completion ratio. The <u>mean tapping rate</u> was calculated in each tapping task as the number of all index finger extensions crossing the lower light beam per second. The <u>tapping variability</u> was calculated as the standard deviation of the normalized movement intervals across a trial. A larger standard deviation means a greater inter-tap-variability of the tapping movements. The movement intervals were obtained as the time intervals between adjacent movement onsets of each trial. Concerning the variation of movement rates between subjects, time intervals were normalized to the maximum interval in a trial. We finally computed the mean tapping variability across trials per task for each subject. The <u>completion ratio</u> was computed as the ratio of index finger extensions reaching the level of the upper photoelectric sensor divided by the total number of index finger extensions.

To estimate the <u>decrement</u>, the trials were first divided into 12 time bins in the slow and the fast tapping task (the first 12s). Decrement was then defined as a decrease of the completion ratios from the first to the twelfth time bin in a trial. We evaluated the decrement by estimating the effect of time bin on the completion ratios in a mixed model (completion ratios ∼ time bin + random(1∣SubjectID)). To compare the decrements in the slow and fast tapping tasks over identical time spans, we also calculated completion ratios for each 1s time bin during the first 12s of slow tapping.

### EEG signal recordings

We recorded high-resolution (24 bit) EEG signals with 64-channels (eego™mylab, ANT Neuro, The Netherlands) at a sampling rate of 2kHz. Vertical electrooculography was also recorded for further removing eye movement components. Bipolar EMG of the first dorsal interosseous (FDI) muscle was recorded from the hand side used to perform repetitive movement tasks. Individual positions of the EEG electrodes and fiducial markers were acquired by a 3D optical digitization system (EEG Pinpoint, Localite, Germany) before EEG recordings. Data were recorded during the 5 min resting period and the subsequent repetitive movement tasks (as mentioned above).

### EEG signal preprocessing

All EEG signal preprocessing procedures were done in the EEGLab Toolbox under a common pipeline. Channels that contained noticeable long-term large artifacts were excluded from the subsequent analysis after raw data of all the channels were demeaned. We applied high pass filtering at 0.5Hz to the data to avoid the slow drifts. Independent component analysis (ICA) in preprocessing was applied to remove the components which contained eye movement artifacts, channel noise, line noise, EKG artifacts, and major muscle artifacts. Artifacts from transitory muscle activities that contaminated the EEG signals were visually detected and marked in the raw data. Subsequently, all data sets were segmented in epochs of 3s duration (−1s to 2s relative to the online mechanical onset). In order to ensure equal treatment of the resting dataset, we randomly generated and inserted 150 fake markers into the original raw datasets, and the information of epochs was saved based on the fake markers. We only used every second movement onset trigger to minimize overlapping of epochs for the fast tapping task. The number of recorded movement cycles varied among different movement tasks, and the least number of repetitions were performed in the pressing task. To obtain a comparable number of movement cycles across conditions, we randomly selected the number of trials in resting, slow tapping, and fast tapping conditions, such that the number of trials matched that derived from the pressing task. The numbers of epochs that were included in the following analysis in each condition are provided in Supplementary Table 2.

### EMG signal processing

The EMG signals were first demeaned, high pass filtered at 0.5Hz, cleared from artifacts, and broadband pass filtered from 5Hz to 200Hz. Notch filters at 50Hz and its harmonics were applied to reduce interference from environmental noise. After rectification, EMG signals were segmented into 3s epochs (−1s to 2s) aligned with the mechanical onset as defined above. EMG signals were smoothed by applying Butterworth low pass filter at 5Hz in 5^th^ order on the rectified signals.^17^ For each subject, the EMG signals were averaged across epochs. Based on the mean EMG signal of each subject, we calculated the EMG slope as the slope between the 25% and 75% percentile of the normalized EMG amplitudes close to the mechanical onset (as shown in Fig. 1E). EMG signals were z-score normalized before computing the EMG slope to diminish individual variability of the EMG amplitudes.

### Region-based source analysis

EEG source analysis was done following the procedures introduced by^18^ and detailed in our previous paper.^3^ In brief, the raw data from EEG sensor signals were projected on the cortical surface employing individual head models and a linearly constrained minimum variance beamformer method^19^ to obtain the source signal in each specific brain region. A combination of principal component analysis (PCA) and independent component analysis (ICA) was applied to reduce noise and obtain signals from components with relatively independent spatiotemporal characteristics in a specific region. Each component could represent a sub-network in that region. In order to eliminate the interference of high-frequency activities that are not in the range of interest and to optimize the calculation of PCA-ICA, before the calculation of the beamformer filter, we applied a 300Hz low pass filter (EEGLab default filter) to the EEG sensor signals recorded under all the conditions. We calculated a common beamformer filter under the merged data of all conditions and applied the PCA-ICA procedure separately to each of the 6 conditions. The PCA-ICA weights were then applied back on the artifacts-marked source signals of the 6 conditions. This study investigated the 4 brain regions that we previously found to show statistically enhanced PAC in patients with Parkinson’s disease compared with controls^3^. These regions were the premotor cortex (PMC), the primary motor cortex (M1), the primary somatosensory cortex (BA3), and the primary somatosensory complex (BA1&2), which were defined with reference to the multi-model parcellation of Glasser et al.^20^. The information regarding the average number of components of each region in patients and controls the 6 conditions are provided in Supplementary Table 3.

### Calculation of movement-related PAC

The raw data of source signals were first filtered into β (13-30Hz) band activities and γ (50-150Hz) band activities. Subsequently, Hilbert transform was applied to extract the phase of the β band and the amplitude of the γ band. Then for each 3s epoch in a component, we computed PAC in successive windows (300ms, corresponding to 4-10 cycles for the β activities) shifted by 50ms time steps. We applied the normalized mean vector length (MVL)^21, 22^ in which the normalized factor is the square root mean of the γ amplitudes in a time window. Since the time window was rather short, which might lead to inaccurate PAC estimation, the z-score of the MVL (zMVL) was calculated using the mean and standard deviation of 200 surrogates created by recombining the instantaneous phase and randomized shuffled amplitudes. zMVL values not larger than 1.96 (equivalent to 95% confidence) were assigned the value 0. Therefore, for each 3s epoch in a component, we had 55 zMVL values with 50ms time resolution.

For each subject, we calculated pairwise zMVL values among ICA component pairs within each region as introduced previously^3^. In brief, we calculated the epochs of the time series of zMVL values among n*n component pairs in a region. We then computed the single time series by computing the weighted average of zMVL values across trials and n*n component. The weights for averaging were defined as the percentage of variance accounted for by each component pair in the region.

#### State-related PAC

To compare PAC across conditions and groups, we computed the mean PAC value by averaging across the 55 time points of 3s-epochs for each subject in each condition.

#### Fluctuations of movement-related PAC

To estimate the fluctuation of PAC over time in different tasks, we calculated the coefficient of variance (CoV) of the movement-related PAC for each task for each subject. CoV was calculated as the ratio of the standard deviation to the mean across 55 time points of averaged PAC for each subject.

#### Dynamic PAC (dynPAC) Estimation

Movement cycles varied between subjects, and movement states within a movement cycle had different durations. Therefore, we first determined in each subject the time points when movement transitions occurred between different movement phases. Data were then aligned with these trigger points to enable averaging dynPAC values regarding movement transitions and assess PAC changes across transitions between movement states. To investigate movement-related PAC dynamics, we subdivided each movement cycle into 5 periods by 4 trigger points derived from the digital kinetic (pressing) or kinematic (tapping) signals recorded during the movement. The trigger points were established individually for each movement cycle in each task. Importantly, to estimate the timing of the transition at the cortex level at the best possible accuracy, we shifted the trigger points by considering the influence of general mechanical, electromechanical delays, and the corticomuscular conduction time across subjects in pressing and slow tapping events relative to the mechanical trigger points. The details for the definition of 4 trigger points mechanically and 4 adjusted trigger points on the cortex level are described in Supplementary Methods.

Therefore, for the pressing task, we have 4 adjusted trigger points indicating: 1) the movement onset; 2) the end of the force build-up; 3) the start of releasing; 4) and the movement offset. Accordingly, the PAC values in movement epochs were grouped into the 5 periods for movements in the pressing task, defined as: P1 (pre-pressing onset period) - during 200ms before the movement onset to the movement onset; P2 (the post-pressing onset period) - the period from the movement onset until the end of the force build-up; P3 (the sustained pressing period) - the period from the end of the force build-up to the start of the force release; P4 (releasing period) - the period from the start of releasing to the movement offset; and P5 (the post-offset period) - the period from the movement offset to 200ms after the movement offset. In the slow tapping task, the 4 trigger points at the cortex level indicated: 1) movement onset. 2) the end of finger extension. 3) the start of finger flexion. 4) the movement offset. Accordingly, the 5 periods were defined as: T1 (pre-extension onset period) – the period from 200ms before the movement onset to movement onset; T2 (post-extension onset period) - the period from the movement onset to the end of finger extension; T3 (extension period) - the period during which the index finger remained extended at or above the upper photoelectric sensor; T4 (index finger flexion period) - the period from the start of finger flexion to the movement offset of the tap; and T5 (post-flexion offset period) - the period from the movement offset to 200ms after the movement offset.

The zMVL values in the single movement cycle were then first grouped in the 5 periods of pressing and slow tapping tasks, respectively. We then averaged the zMVL values in each of the periods across trials for each subject.

### Power spectral density

The power spectral density (PSD) was calculated by the Welch method implemented in Matlab. In order to be comparable with the movement-related PAC, the PSD was calculated in 300ms shifted window with 50ms time steps in 3s-epochs (Hann window, frequency resolution 1Hz) of all conditions. Then, the PSD was transformed in base 10 logarithmic power for group comparison and presenting. The power was normalized by subtracting the mean power across all time points and trials from 4 to 300Hz (excluding 50Hz and its harmonics) to account for the inter-subject variability.

### Statistical test

All analyses were performed in the brain regions contralateral to the hand side on which the subject performed the movement tasks. We mainly applied ANOVA tests to examine the main and interaction effects of the factors we have obtained in the analysis. Since our data were not always Gaussian distributed and of equal variance, we applied a non-parametric ANOVA provided by the ARTool package (Aligned Rank Transform for non-parametric factorial ANOVAs).^23^ This package applies the aligned rank transform to the responses of each main or interaction effect in the designed model and then runs a factorial ANOVA (type III Wald F tests with Kenward-Roger degree of freedom) on the transformed data. For post-hoc tests, we applied a non-parametric Wilcoxon rank-sum test for between-group comparisons and Wilcoxon sign-rank test for within-group comparisons across conditions and movement transitions. We also computed Spearman correlations among PAC, performance parameters, and clinical severity scores. The false discovery rate (FDR) was applied in multiple tests to avoid type I errors in null hypothesis testing.

## Data availability

Personal data are protected by data privacy statements signed by all subjects. The data can be made available upon specific request taking into account the opinion of the local data privacy board.

## Results

### Behavioral analysis

The behavioral analysis provided evidence for slowed motor performance in the patients that varied by tasks and observed parameters (Fig. 2). In the pressing task, patients and controls performed press-release actions at a similar rate (patients, 0.33±0.08 /s; controls, 0.31±0.09 /s; p=0.633; Fig. 2A-i). The maximum EMG amplitude did not differ between patients and controls (p=0.684, Fig 2A-ii). Likewise, the EMG slopes regarding either force build-up or releasing were similar between patients and controls (rank-sum tests, force build-up, p=0.527, Fig 2A-iii; releasing, p>0.99).

**Figure 2.**
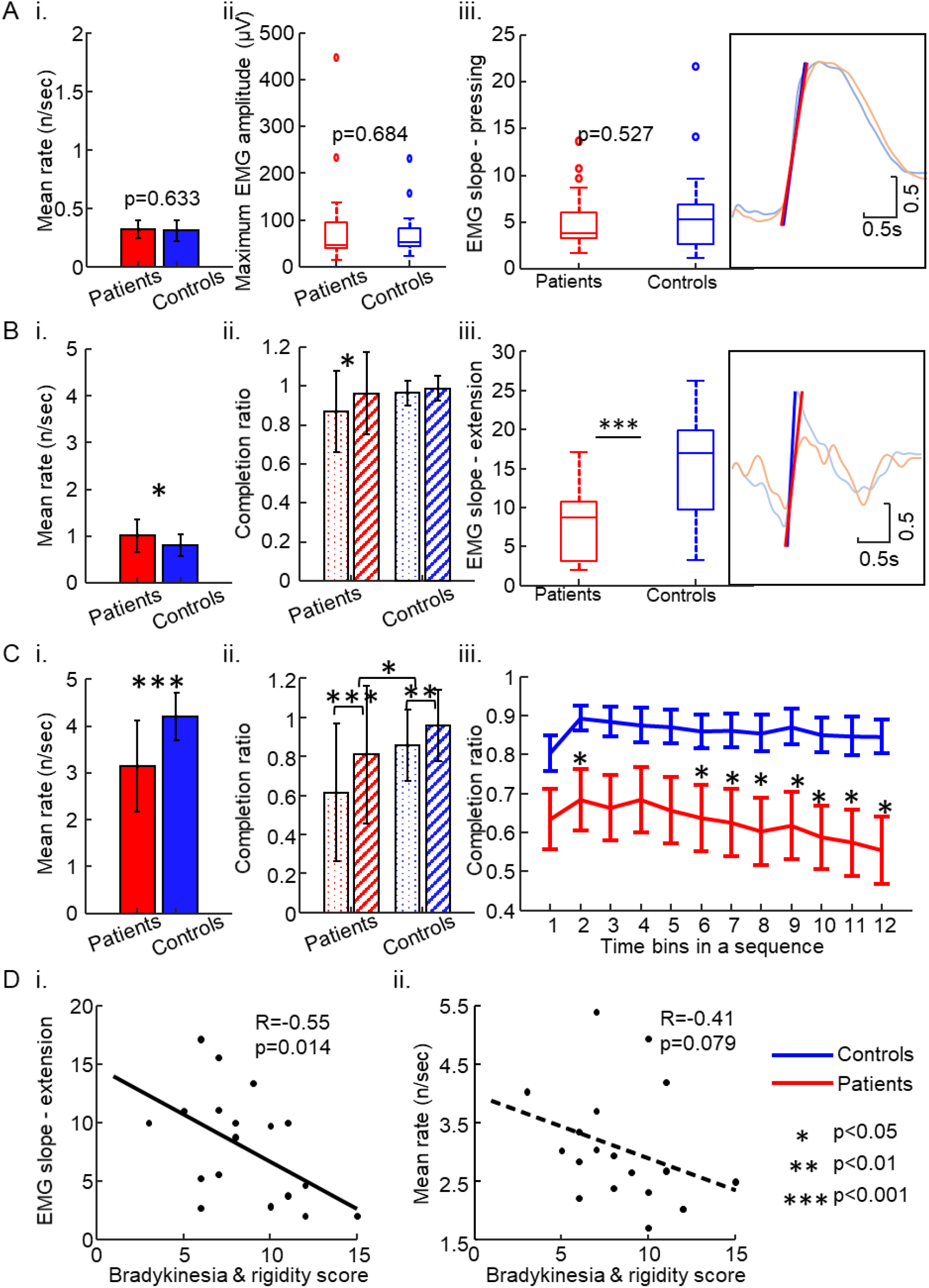
Selected performance parameters of the repetitive movement tasks. A. Pressing: (i) rate of repetitive pressing actions, (ii) maximum amplitude of EMG recorded from FDI muscle during pressing, (iii) slope of EMG activity in FDI muscle during the build-up of pressing force. Inset: averaged EMG curves of patients (red) and controls (blue). B. Slow tapping: (i) rate of repetitive slow tapping actions, (ii) completion ratio of index finger extensions meeting the upper amplitude criterion (dotted: without feedback, hatched: with feedback), (iii) slope of EMG activity in FDI muscle upon index finger extension. Inset: averaged EMG signals of patients (red) and controls (blue). Note less steep EMG slope in patients. C. Fast tapping: (i) rate of repetitive fast tapping actions, (ii) rate of index finger extensions meeting the upper amplitude criterion (no visual feedback about the level of index finger extension), (iii) decline of fraction of extension movements meeting the upper amplitude criterion across sequential 1s time bins. Values denote mean +/- s.e.m., D. Spearman rank correlations between (left) EMG slope of patients at tapping onset or (right) mean tapping rate with hemi-score for bradykinesia and rigidity of the MDS-UPDRS III for the more affected side.

In the tapping tasks, we applied a two-way mixed ANOVA with the factors Group (2 levels: patients and controls) and Feedback (2 levels: with or without feedback) on the three performance parameters (tapping rate, tapping variability, and completion ratio). In slow tapping, the tapping rate was slightly higher in patients than in controls (main effect: Group, F(1,37)=4.68, p=0.037, Fig 2B-i). With visual feedback, the tapping rate increased in both groups (main effect: Feedback, F(1,37)=14.63, p<0.001), probably through an effect of pacing by the visual feedback. The tapping variability did not differ between patients and controls (main effect: Group, F(1,37)=0.19, p=0.667), and the group comparison was not affected by feedback (main effect: Feedback, F(1,37)=0.11, p=0.74). We found a significant interaction effect between Group and Feedback on the completion ratio (F(1,37)=8.91, p=0.005, Fig. 2B-ii). Post-hoc testing revealed that, although no significant differences were found between patients and controls in either the without-feedback (p=0.115) or the with-feedback condition (p=0.220), the completion ratio increased with visual feedback in patients (p=0.005), but not in controls (p=0.081). The decrement of tapping amplitude, as indexed by the decrease of completion ratio over a trial duration, was evaluated for tapping without visual feedback. Mixed ANOVA analysis on the completion ratio in the sequence of 12 time-bins within a 30s trial revealed an interaction between Group and Time Bins in the slow tapping task (F(11,407)=7.19, p<0.001). However, completion ratios did not differ between patients and controls in any of the time bins after correction (all p values>0.2; FDR-corrected). Subsequently, we computed the mixed-effects model ‘Completion ratio ∼ Time Bin + random(Subjects)’, separately for patients and controls. In slow tapping, the effect of Time Bin on the completion ratio was significant in patients, due to a decline over time (t(226)=-4.69, p<0.001), and in controls, in whom the completion ratio even slightly increased over time (t(198)=3.87, p<0.001). When we considered only the first 12s of slow tapping, in order to facilitate comparison with the fast tapping task (see below), the decrease in the completion ratio over a trial was not statistically significant in patients (t(226)=-1.58, p=0.115). The EMG slope at tapping onset (index finger extension) was significantly lower in patients compared with controls (p<0.001, Fig. 2B-iii).

In the fast tapping tasks, the tapping rate was lower in patients (main effect: Group, F(1,37)=18.22, p<0.001, Fig. 2C-i) and faster in both groups with feedback (main effect: Feedback, F(1,37)=9.46, p=0.004). There was no interaction effect between Group and Feedback (F(1,37)=0.91, p=0.34). Tapping variability was higher during fast tapping in patients compared with that of controls (main effect: Group, F(1,37)=14.33, p<0.001). Visual feedback had no effects on the tapping variability (main effect: Feedback, F(1,37)=0.68, p=0.416). For completion ratio, ANOVA revealed a significant interaction effect between Group and Feedback (F(1,37)=8.65, p=0.006, Fig. 2C-ii). Post-hoc tests revealed that the completion ratio was lower in patients than in controls, both without feedback (p=0.021) and with feedback (p=0.013). With visual feedback, both patients (p<0.001) and controls (p=0.004) improved their tapping performance by meeting the upper height criterion more frequently. Patients improved more than controls. Regarding decrement, the mixed ANOVA analysis on the completion ratio in the sequence of the first 12s within a trial revealed an interaction between Group and Time Bin (F(11,407)=3.05, p<0.001, Fig 2C-iii). Comparing patients and controls in the sequence of 12 time bins within a trial revealed significant differences between the two groups (Figure 2C-iii). Furthermore, evaluating the decrease of the completion ratio along the sequence of time points, we found a significant effect of Time Bin on the decrease of the completion ratio in the patients (t(226)=-5.45, p<0.001).

In summary, whereas in pressing all performance metrics of patients were similar to those of the control subjects, in slow tapping the evidence for abnormal slowness was limited to the speed of muscle recruitment, and the tapping rate was unaffected. In fast tapping, the tapping rate was reduced, and decrement was evident.

We explored the relationship between various abnormal performance metrics in the tapping tasks and the clinical severity of motor impairment in patients as indexed by the hemi-body bradykinesia and rigidity scores from MDS–UPDRS III. Of all parameters, only the EMG slope at tapping onset in slow tapping was significantly correlated with the hemi-body bradykinesia and rigidity scores (Fig 2D-i, p=0.014, R=-0.55). There was a tendency of a negative correlation between the fast tapping rate and the bradykinesia and rigidity scores (Fig 2D-ii, p=0.079, R=-0.41). The tapping variability did not show any significant correlation with clinical severity. Moreover, there was no correlation between the clinical score and the decrement computed as the difference of the completion ratios between the 1st and 12th time bins in either the slow tapping or the fast tapping task.

### Similar state-related PAC during tapping tasks between patients with Parkinson’s disease and controls

We examined whether PAC averaged over the whole movement period of each different condition (“state-related PAC”) was modulated differently by different tasks in patients and controls. To reduce the number of factors, we first established that the visual feedback did not modulate the strength of state-related PAC (3-way ANOVA with factors Group (patient, controls), Region (4 levels) and Feedback (FB+, FB-) for slow tapping task and fast tapping task). The factor Feedback showed neither interaction nor main effects in either the slow (F(1,37)=0.26, p=0.616) or the fast tapping tasks (F(1,37)=0.04, p=0.840). Therefore, in the following analysis, we combined the conditions with and without feedback within the slow and fast tapping tasks. A three-way (Group, Task, Region) mixed ANOVA performed on the zMVL values averaged over the entire 3s-epochs suggested that state-related PAC was modulated differently in patients and controls by the tasks (Group x Task; F(3,111)=4.69, p=0.004; Fig. 3). Post-hoc rank-sum testing revealed enhanced PAC in patients compared to controls in the resting state (p=0.016), in agreement with previous findings (Gong et al., 2021). PAC was also enhanced in patients in the pressing task (p=0.012), but not in any of the tapping tasks (p values>0.5). Post-hoc testing also showed that state-related PAC in patients was reduced during all active movement tasks compared to rest (resting vs. pressing, p=0.033; resting vs. slow tapping, p=0.010; resting vs. fast tapping, p<0.001). In controls, we found no significant reduction of state-related PAC during pressing (p=0.296), during slow tapping (p=0.654) or during fast tapping (p=0.135) compared to the resting state. The ANOVA also revealed that PAC differences among the conditions were affected by the motor control area (Task x Region; F(9,333)=2.64, p=0.006). However, since we found no interaction effects among the three factors (Task x Region x Group; F(9,333)=0.40, p=0.933), the effect of brain regions was unlikely to affect the differences between groups.

**Figure 3.**
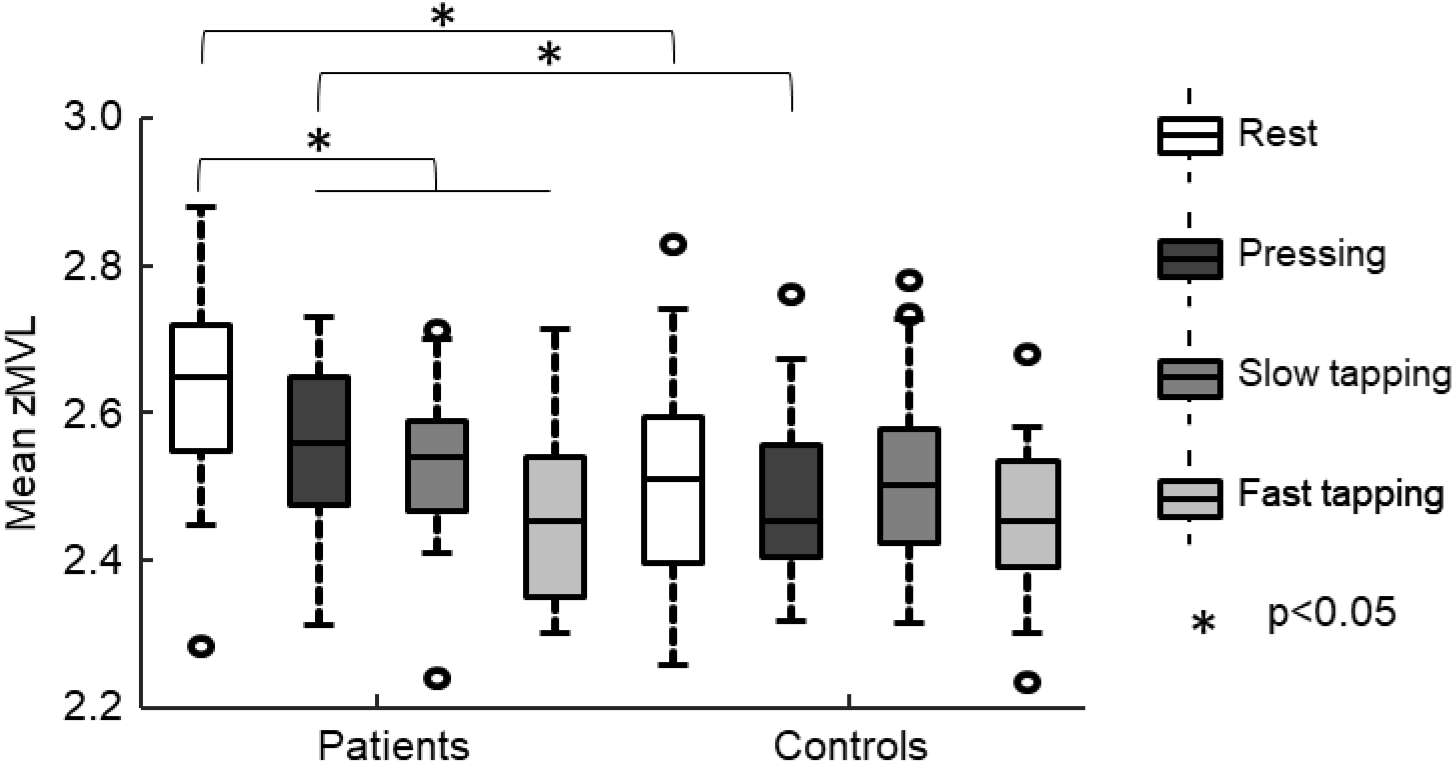
State-related PAC for the different tasks, averaged across 4 regions. A three-way non-parametric ANOVA showed significant interaction effects (designated by an asterisk) between Group and Tasks. Post-hoc tests of the Disease*Task interaction effect showed significant differences (designated by an asterisk) between the resting state and all 3 movement tasks in the patients. Note that state-related PAC differed between patients and controls in resting state and during pressing.

The above findings showed differences between the resting state and movement-related state-related PAC, especially in patients. Among the movement tasks, state-related PAC differed between patients and controls only in the pressing task, where motor behavior was similar between patients and controls. In contrast, state-related PAC was remarkably similar between patients and controls in the tapping tasks, where motor behavior was different. Because state-related PAC was derived from EEG recorded during active movement, abnormal enhancement of PAC per se is unlikely to be directly related to motor impairment.

### Dynamics of PAC during transitions between different movement states

We then considered the possibility that the dynamic modulation of PAC might be more directly related to the underlying pathophysiology of motor impairment than its absolute level. We first visualized dynamic PAC and EMG activity recorded from the FDI muscle, aligned with the online mechanical movement onset (Fig. 4A). Alignment to movement onset confirmed that in patients, PAC was generally reduced in all movement tasks compared to the resting state, as shown in the previous paragraph. Additionally, it became evident that in controls, the PAC values in the pressing and slow movement tasks were markedly modulated along the movement cycle. In contrast, this modulation was considerably less pronounced in the resting state and during fast tapping. For pressing and slow tapping, PAC rapidly and markedly declined from a brief peak before rebounding again around movement onset. Modulation appeared to be less marked in patients.

**Figure 4.**
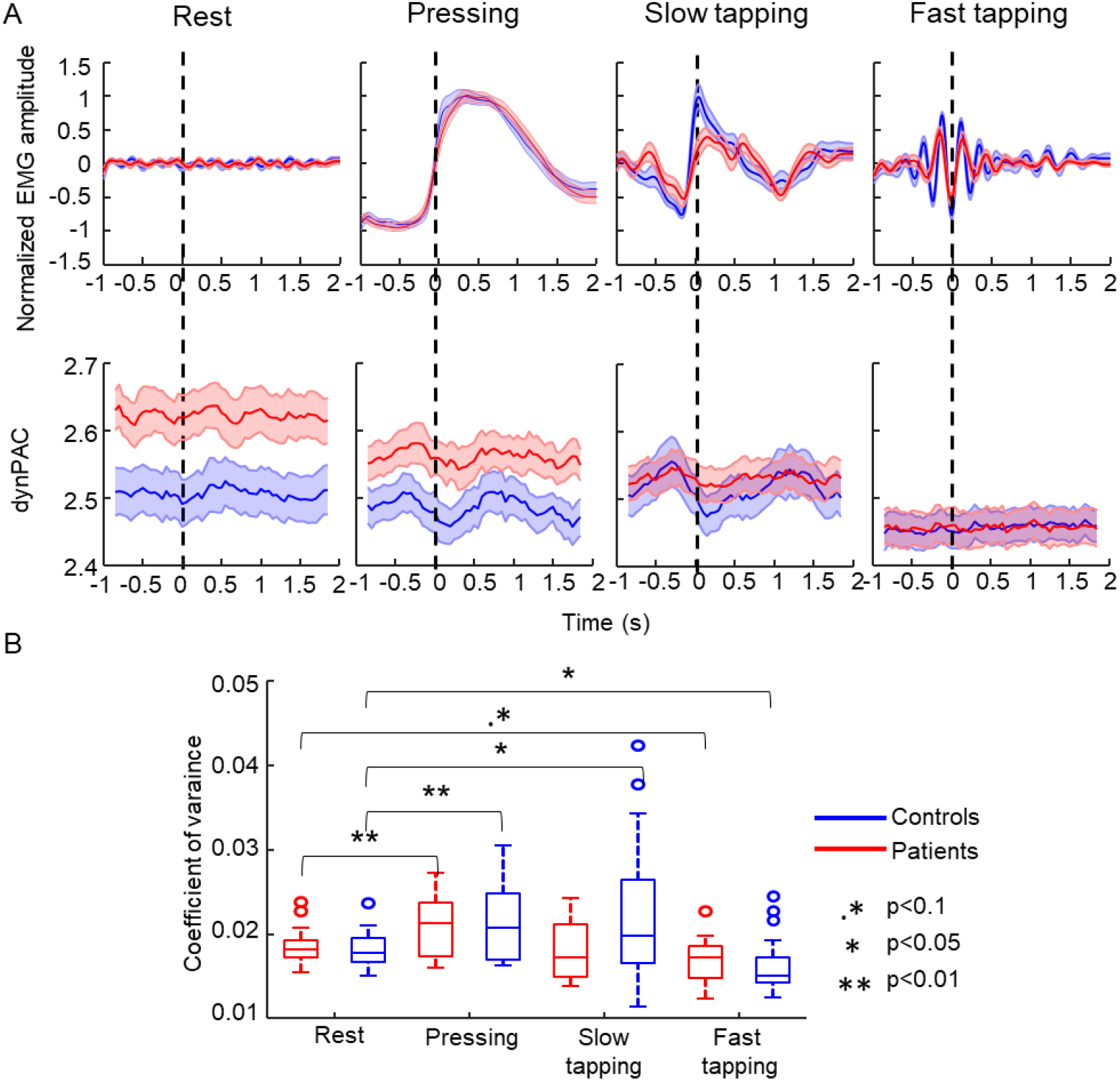
Fluctuation of PAC in the 3s time series. A. PAC dynamics in the 4 conditions. Upper panel, rectified EMG (mean ± SE), recorded from first dorsal interosseus muscle, averaged after alignment to arbitrary time points (rest) or to movement onset (pressing, slow and fast tapping). Lower panel, PAC (mean ± SE) averaged after alignment to arbitrary time points (rest) or to online mechanical onset (pressing, slow and fast tapping). B. Coefficient of variance of PAC across the 4 conditions.

To statistically assess the degree of fluctuation of movement-related PAC for each task quantitatively, we first computed the CoV across the time series. A two-way mixed ANOVA test (CoV ∼ Group * Task) showed significant interaction (F(3,111)=3.07, p=0.031, Fig. 4B). In pressing, post-hoc testing revealed that the fluctuation was stronger for both patients (sign-rank, p=0.005) and controls (p=0.006) compared with the resting state. In slow tapping, the fluctuation was larger than in the resting state in controls (sign-rank, p=0.033), but not in patients (p=0.872). In fast tapping the tapping rate (>4 Hz) implies a duration of each full movement cycle of less than 250ms. Since the resolution of PAC calculation does not permit assessment of modulation across very short movement cycles, reductions of fluctuation of PAC across a movement cycle (sign-rank, controls, p=0.044; patients, p=0.084) must be interpreted with caution. Direct comparisons of CoV between patients and controls did not reveal any differences in the resting state (rank-sum, p=0.567), pressing task (p=0.967), and fast tapping task (p=0.244), but a marginal difference in the slow tapping task (p=0.070). The larger time-series variance of PAC in pressing and slow tapping tasks compared with the resting state may hint at the possibility that PAC may be modulated at particular movement events. Therefore, we subsequently investigated the modulation of movement-related PAC across transitions between different movement states in the pressing task and the slow tapping task (“dynamic PAC”).

We investigated the dynamics of PAC in more detail by looking at the modulation of PAC between 5 periods separated by 4 trigger points. The dynamic trigger points were calculated using 4 kinetic (pressing) or kinematic (tapping) events along the movement cycle (Fig. 5A), taking into account mechanical delays, electromechanical delays, and the corticomuscular conduction time to estimate the timing of the transition times between motor states at the level of the cortex at the best possible accuracy (Fig. 5B&D). The details for the definition of the four adjusted trigger points are described in the method section.

**Figure 5.**
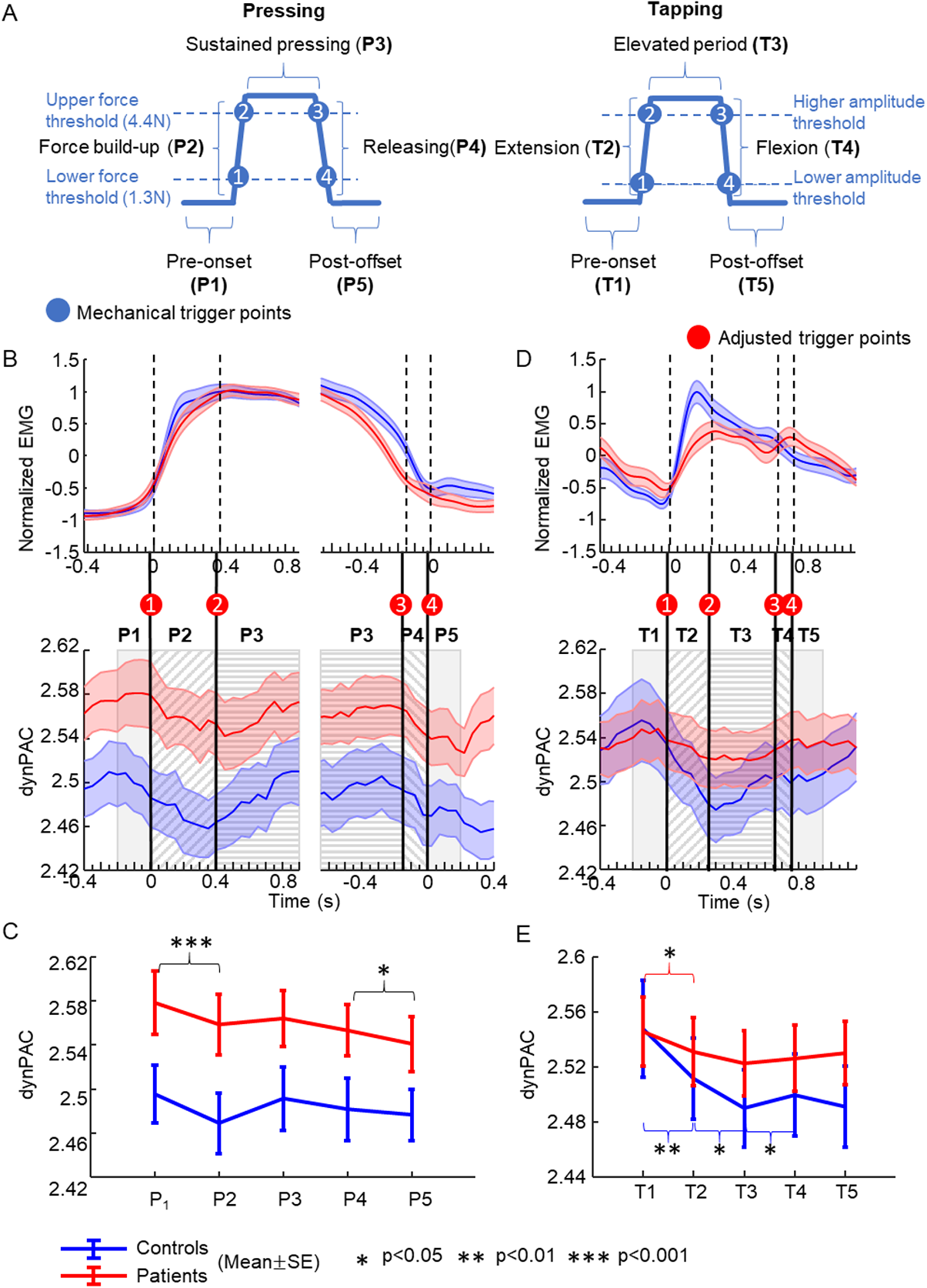
PAC dynamics across movement transitions. A. Definition of 5 periods of movement transitions based on 4 trigger points in the pressing task (left) and the slow tapping task (right). The trigger points were first established using 4 kinetic (force threshold for pressing) or kinematic (amplitude criterion for tapping) events along the movement cycle. The adjusted trigger points (displayed in B&D) were then determined by further considering the mechanical delays, electromechanical delays and the corticomuscular conduction time to estimate the timing of the transition times at the level of the cortex. These 4 trigger points designated movement transitions separating five movement periods. B. Pressing task. Dynamic PAC as resulting from averaging after alignment with the adjusted trigger points on the (left) onset (#1) and (right) offset (#4). The timing of the other 2 trigger points (left: trigger point #2, right: trigger point #3) was determined based on the averaged durations of periods (left: P2 and P3, right: P3 and P4) across subjects. Note similar PAC modulation pattern in pressing onset and offset. C. Average movement-related dynamic PAC in each of the 5 periods of a single pressing cycle in patients and controls. Patients displayed enhanced PAC in all periods compared with controls. Note similar modulation patterns in patients and controls. D. Tapping task. Dynamic PAC as resulting from averaging across subjects after alignment with the adjusted trigger point #1. The timing of the other 3 adjusted trigger points was based on the average duration of the following 3 periods (T2, T3, T4) across subjects. E. Average movement-related dynamic PAC in each of the 5 periods of a single tapping cycle in patients and controls. Patients displayed markedly less modulation of PAC than controls.

In the pressing task, PAC was markedly modulated around movement transitions. Fig. 5B illustrates the averaged dynamics of PAC across the first 3 periods as resulting from aligning data with the adjusted trigger point #1 (defining the transition between pre-movement onset and force build-up, left panel), and the last 3 periods as resulting from aligning data with the adjusted trigger point #4 (defining the transition between force release and pressing offset, right panel). As shown in Fig. 5B, close to the onset of pressing, PAC appeared to decrease from a brief maximum, reaching a minimum before rising again (also evident in Fig. 4). Alignment with the transition at the offset of releasing revealed a PAC motif similar to that at the onset of pressing (Fig. 5B, right panel). Two-way mixed ANOVA on Group and Period showed significant main effects for the factors Group (F(1,37)=5.57, p=0.024) and Period (F(4,148)=4.72, p=0.001), whereas there was no interaction effect between the two factors (F(4,148)=1.08, p=0.369). As shown in Fig. 5C, post-hoc testing indicated that, across subjects, dynPAC decreased significantly during the build-up of the pressing force compared with the pre-onset period (P1 vs. P2, p<0.001). Then, the PAC value rebounded during maintained pressing (P2 vs. P3, p=0.070) at marginal effects. DynPAC showed a tendency to decrease from P3 to P4 (P3 vs. P4, p=0.118), while the PAC values significantly decreased from force releasing to the period after pressing offset (P4 vs. P5, p=0.034). This finding indicated that dynPAC was modulated across movement transitions before force build-up and after releasing actions. Because there was no Group x Period interaction effect, PAC was not differently modulated between patients and controls.

We also investigated PAC dynamics in slow tapping movement cycles in a manner similar to the pressing task. Since the presence or absence of color feedback had no significant effects on the PAC values in slow tapping, the evaluation of PAC dynamics was based on combining the two conditions. Similar to the pressing task, we found marked modulation of dynPAC in the slow tapping task. As shown in Figure 5D, PAC declined from a brief maximum before index finger extension onset to post extension onset in controls. After reaching a minimum when the index finger was above the higher amplitude threshold, PAC started to rebound and increased toward the post-offset period. Although the modulation pattern was generally similar in patients, it was flattened compared to controls (also evident in Fig. 4). A two-way mixed ANOVA of dynPAC values on the factors of Group and Period revealed a significant two-way interaction between Group and Period (F(4,148)=3.59, p=0.008). This finding indicated that PAC was modulated differently in patients and controls. Post-hoc testing revealed that while dynPAC decreased from T1 to T2 in both patients (sign-rank, p=0.016) and controls (sign-rank, p=0.006), the decrease of PAC associated with index finger extension (T2) was smaller in patients than in controls, as shown in Fig. 5E. Then there was a continuous decrease of PAC from T2 to T3 (sign-rank, p=0.005) followed by a rebound from T3 to T4 (sign-rank, p=0.021) in controls, while the modulation was less pronounced in patients (sign-rank, T2-T3: p=0.260, T3-T4: p=0.260). From the finger-flexion period to the post-flexion offset period, no significant PAC increase was found in either controls (sign-rank, p=0.126) or patients (sign-rank, p=0.520). Notably, the absolute PAC values did not differ between patients and controls in any of the 5 periods (rank-sum, all p >0.2). This finding showed less PAC modulation in patients during selected periods of the tapping cycle than in controls during slow tapping movement. We subsequently tested the hypothesis that the magnitude of the PAC change around movement onset determines the ability to recruit muscles engaged in the tapping rapidly and thus may contribute to the motor impairment in slow tapping performance in patients. Although the EMG slope was correlated with the PAC change between T1 and T2 in patients, this correlation was lost when the computation of PAC was corrected for the shorter averaged duration of T2 across subjects. EMG slope and PAC change were not significantly correlated in controls (R=-0.12, p=0.601).

### Relationship between movement-related PAC dynamics and β power dynamics

It is well established^24-26^ that spectral power in the β-frequency band decreases during the initiation and execution of a movement. In the present study, we also found β power to be reduced at movement onset and during movement in both patients and controls (Fig. 6 A&B, left panels). Because the strength of β power affects the estimation of phases in the calculation of PAC, we aimed to investigate to which degree modulation of PAC by movement transitions was a consequence of associated changes in β power. We assessed the modulation of β power in the 5 periods of a movement cycle in both the pressing and the slow tapping task (Fig. 6 A, B, right panels). The modulation pattern of β power appeared to be similar to the modulation pattern of PAC in both tasks as reported above. Two-way mixed ANOVA with Group and Period was applied on the β power in pressing task and in slow tapping tasks. ANOVA in the pressing task revealed only a main effect of Period (F(4,148)=13.18, p<0.001), while the ANOVA in the slow tapping task revealed a significant interaction of Group and Period (F(4,148)=3.98, p=0.004). This finding raises the question of whether the transient modulation of β power primarily drove PAC modulation during movement. We did not find a significant correlation between the absolute PAC and β power values in any of the periods in either the pressing task or the slow tapping task (p values > 0.3). Two examples of scatter plots between absolute power and PAC in P1 (T1) of pressing (slow tapping) are displayed in Fig. 6C. Additionally, we performed a correlation analysis between the PAC differences and the β power differences of each two adjacent periods. At movement onset, the two parameters were not significantly correlated in the pressing task (R=0.13, p=0.429), whereas they were significantly correlated in the slow tapping task (R=0.50, p=0.003). The correlation results for differences between any two adjacent periods are presented in Table1, which showed no consistent relationship between β power change and PAC change during the movement. The above findings suggest that the movement-related dynPAC modulation does not generally reflect movement-related power dynamics. However, the fact that derivatives of the two variables were correlated at slow tapping onset could still have pathophysiological significance.

**Figure 6.**
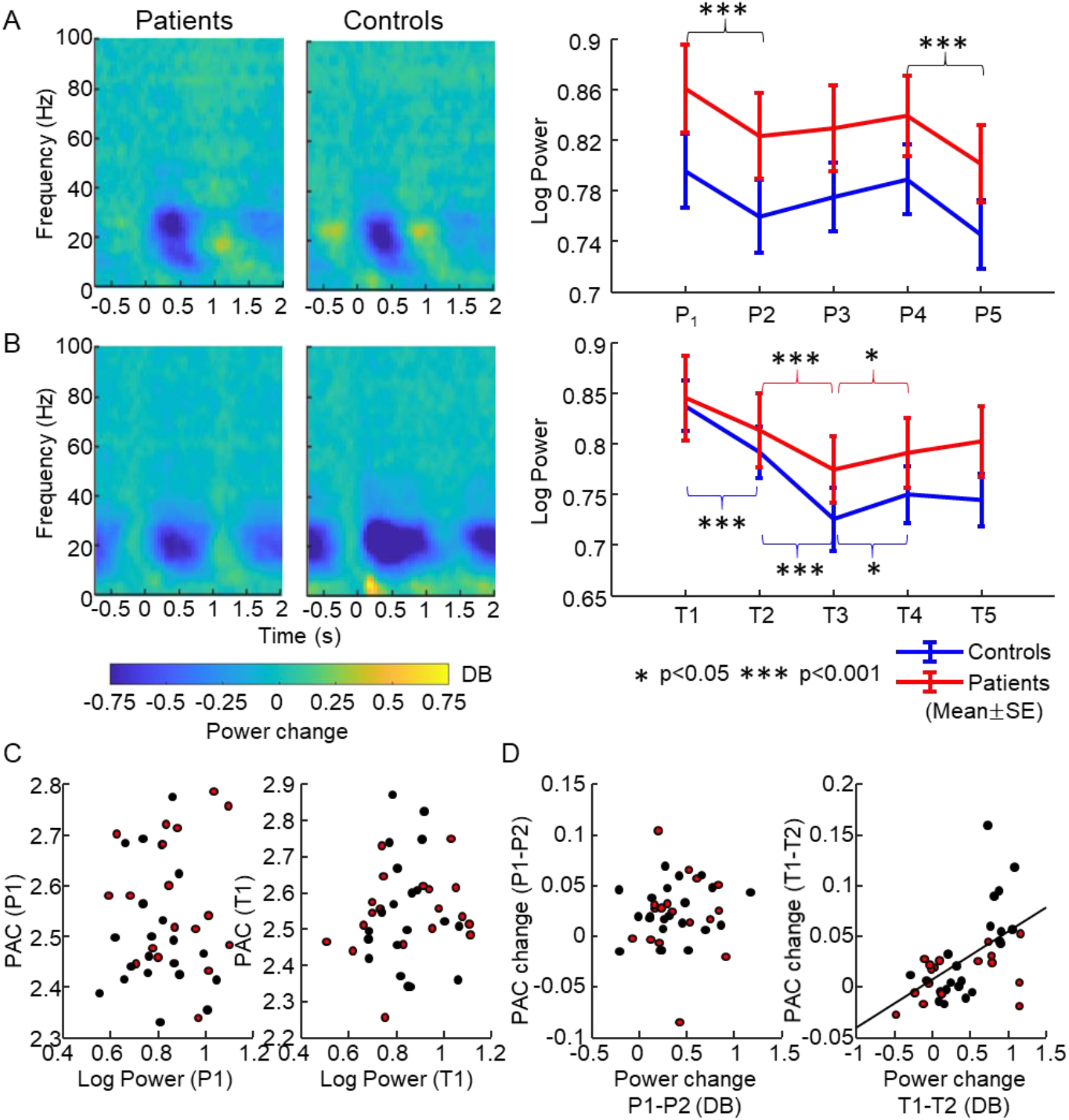
Relationship between movement-related dynPAC and β power. A. Dynamics of β power in the pressing task. Left, time-frequency spectrogram. Note the reduction of β power after the onset of the pressing in both patients and controls. Right, dynamics of β power across the 5 phases of the pressing movement cycle. Significant modulations were marked by an asterisk. B. Dynamics of β power in the slow tapping task. Left, time-frequency metric plots. Right, dynamics of β power across the 5 phases of the slow tapping movement cycle. Significant modulations were marked by an asterisk. C. Scatter plots of the relationship between the absolute strength of PAC and β power in the pre-onset phase. D. Scatter plots of the relationship between PAC change and power change from the first to the second period of the movement cycle. Left, pressing task. Right, slow tapping task.

## Discussion

The present study examined whether and how β-γ phase-amplitude coupling (PAC) of oscillatory neuronal activity in the cortex, as recorded by surface EEG signals during movements, is related to the motor impairment in Parkinson’s disease.

Although previous studies have found that enhanced β-γ PAC at rest re-normalized^7^ or remained enhanced^1, 11^ during movements, this question has remained open - in part because it remained unclear in these studies whether movement-related PAC was derived from signals recorded during the execution of kinematically normal or abnormal movements.^1, 7, 11^ The present study addressed this ambiguity by comparing different types of movements, with different probabilities of revealing Parkinsonian motor impairment, in the same cohort of patients. In a pressing task consisting of self-paced repetitive pressing and releasing actions, we found that movement-related PAC was persistently enhanced in patients compared with controls in 4 cortical motor control areas, namely PMC, M1, BA3, BA1&2. However, the behavioral performance in that task, as indicated by the rate of self-paced movements and by the slope of the EMG in a muscle contributing to the pressing, did not differ between patients and controls. In contrast, there was evidence of abnormal motor behavior in patients in repetitive tapping tasks, but no differences of the movement-related PAC level were found between patients and controls. Behaviorally, the impairment of patients with Parkinson’s disease was evident in the reduced EMG slope, slowness of the movement rate in the fast tapping task, and in the decrement of the amplitude in both slow and fast tapping, which is consistent with previous reports.^14, 27, 28^ These results show that, although movement-related PAC differed between patients and controls depending on the type of repetitive movements, abnormal enhancement of state-related PAC in movement states is not directly related to the motor impairment of Parkinson’s disease.

We found that PAC decreased around movement onset in both patients and controls. This reduction of PAC during movement is in line with studies where PAC was derived from oscillatory signals recorded from STN, globus pallidus, and the primary motor cortex.^1, 7, 10, 11^ It has long been known that voluntary motor activity is accompanied also by event-related β-power desynchronization in the cortex.^26, 29, 30^ However, it was shown that the magnitude of PAC is not related to the absolute β power either in the resting state^2, 6^ or in the movement states as shown in our study. Likewise, the dynamics of PAC during movement were not consistently associated with the β power change shown in our study and in previous literature.^10^ Therefore, the dynamic modulation of β-γ PAC during repetitive voluntary movements may encode an essential component of the motor command that is different from the mechanism underlying event-related desynchronization.

Further insight into the role of PAC in motor control was provided by the detailed analysis of its dynamical modulation across a movement cycle (dynPAC). When controls performed the pressing task, PAC exhibited a brief peak followed by a decline during the initial pressing phase and a subsequent rebound while the index finger still maintained a constant press. Of note, during the initial releasing phase, dynPAC showed a similar pattern (decrease following a brief peak). Also, in the slow tapping tasks, a similar pattern with a brief peak followed by a decrease of PAC was present around the onset of the finger extension, followed by a rebound. These findings in healthy controls suggest that PAC decrease is not merely associated with initiating a movement. Rather, there appears to be a characteristic PAC motif (brief peak – decrease – rebound) that signals a change in movement states. This phenomenon resembles the preparatory neuronal activity in the dynamical systems theory of motor control^31^. According to this theory, preparatory activity brings the dynamical state of the neuronal population through state-space rotations to an initial value. This process, which is characterized by brief cortical oscillatory activity^32^, ensures that muscle activity can be generated efficiently for all types of movements.^31, 32^ If dynPAC reflects normal preparatory activity, then it is perhaps not surprising that movement-related PAC was found to be similar in patients with Parkinson’s diseases and with essential tremor^10^, especially in the absence of kinematic differences between both patient groups.

In patients with Parkinson’s disease, we found that dynPAC was abnormal in slow tapping. While PAC values attained similar levels before initiating the tapping movements, the subsequent decrease was smaller and the later rebound less marked than in controls. In contrast, during pressing, dynPAC modulation was similar in patients and in controls. These findings appear to be the first to report that abnormal PAC during movement is associated with concurrent abnormal motor performance in Parkinson’s disease. If the above hypothesis of dynPAC as a marker for a preparatory movement state is accepted, patients with Parkinson’s disease may suffer from a defective evolution of a neuronal population dynamic from the preparatory state to overt movement generation. While PAC would be a physiological phenomenon at the preparation of movements, its persistence into the unfolding movement would interfere with the proper execution of the movement. Although abnormal dynPAC modulation was associated with slowed muscle recruitment during onset of slow tapping, the PAC change did not correlate with the magnitude of the EMG slope suggesting a complex and non-linear relationship between dynPAC and the build-up of corticospinal neuronal activity. Interestingly, studies probing cortical physiology in the preparatory phase of voluntary movements have provided similar evidence to suggest that bradykinesia does not result from a single deficient physiological mechanism such as the ability to release from ongoing inhibition^33, 34^ but reflects a more complex circuit abnormality.^35^ Notably, preparatory activity in dynamical systems theory of motor control is sensitive to timing events supporting motor transitions^36^, but does not reflect specific movement features (e.g., direction, force, velocity), nor does it simply represent the release from inhibition of a motor program.^37^

The PAC change was significantly correlated with the change of spectral power in the β-frequency band around tapping movement onset, which was characterized by abnormal EMG recruitment. Although this correlation was not specific to slow tapping onset, this finding may suggest that β-frequency activity could at certain critical times have pathophysiological significance for dynPAC. One might speculate that the higher frequency or duration of β bursts in STN^38^ around the onset of bradykinetic movements allows more time to stabilize PAC abnormally. However, as there was no relationship with the absolute β power the significance of this finding is still uncertain.

The question arises as to why PAC is elevated at rest and why its magnitude is associated with Parkinsonian motor impairment as reported in previous literature.^3, 6, 7^ One possibility would be that PAC at rest, or any state-related PAC, is not mechanistically related to the dynPAC abnormality found during slow tapping. Animal experiments may reveal whether state-related PAC and dynPAC could map onto deficient tonic and phasic dopamine activities^39, 40^, respectively, which may be tied to deficits in the invigoration of movements.^39, 41^ If, however, both PAC phenomena are based on a common mechanism, they could reflect processes of movement preparation. In this scenario, on the one hand the increase of dynPAC would indicate a preparatory state during movement dynamics, and its reduced attenuation at movement transitions would indicate a spillover of the preparatory state into the unfolding movement. On the other hand, enhancement of PAC at rest could reflect the abnormal, or abnormally frequent, generation of brief cortical states resembling preparatory population activity in the absence of an intention to move. In this way, both the enhanced resting PAC and the reduced dynPAC modulation could be caused by the dysfunction of a single (subcortical) mechanism that controls or regulates the generation of cross-frequency couplings in cortical microcircuits. However, in the absence of direct evidence, the nature of the link between abnormally enhanced PAC at rest and abnormal modulation of dynPAC must remain a topic for future investigations.

### Limitations

Although self-paced tapping movements may be considered relatively elementary movements and although they reflected the motor impairment of Parkinson’s disease in our study, it is possible that different mechanisms underlie performance impairment in other types of movement. Therefore, future studies need to explore abnormal cross-frequency coupling more comprehensively during more variable motor behaviors. The resolution of the analysis of the dynamics of PAC across the movement cycle is constrained by the minimum number of oscillatory cycles required to compute PAC which, in turn, depends on the involved oscillation frequencies. Therefore, dynamic modulation of PAC cannot be resolved during fast repetitive movements. In addition, separation of the movement cycles into discrete periods may not apply to everyday behavior, which in many instances is more appropriately conceived of as a continuous action with no discrete transitions. It remains to be studied whether concepts derived from analyzing movement periods translate into the control of continuous movements. As noted before^3^, patients with marked resting tremor were excluded, and all recordings were done in patients with early to moderate disease stages. Therefore, it remains unclear how generalizable our findings are to tremulous or more severe Parkinson’s disease phenotypes.

In conclusion, the present study has provided evidence to demonstrate that PAC may serve a role in normal motor behavior, where it appears to indicate a preparatory state of the motor system. Association of abnormal PAC dynamics with bradykinesia is compatible with the hypothesis that deficient regulation of PAC is causally involved in the pathophysiology of Parkinsonian motor impairment.

## Acknowledgement

We thank all patients and healthy volunteers who participated in our study. We also thank Alhuda Dabbagh for assisting in data collection.

## Abbreviations

BA: Broadman area
CoV: coefficient of variance
dynPAC: Dynamic PAC
ECoG: electrocorticography
ICA: independent component analysis
LFP: local field potentials
MDS-UPDRS III: part III of the Movement Disorder Society Unified Parkinson’s Disease Rating Scale
M1: primary motor cortex
PAC: phase-amplitude coupling
PMC: premotor cortex
PSD: power spectrum density
STN: subthalamic nucleus

