## Supplementary Material for "Cross-frequency phase-amplitude coupling in repetitive movements in patients with Parkinson’s disease"

#### Supplementary methods

##### 1.1 The definition of mechanical trigger points

In order to investigate the dynamics of brain activities during transitions between movement states, we first determined in each subject when movement transitions occurred between different movement phases. Therefore, we separated the movement into five periods separated by four trigger points derived from the digital kinetic (pressing) or kinematic (tapping) signals real-time recorded during the movement for each individual. Trigger points were established individually for each movement cycle in each task. Since the movement transient is too quick during the fast tapping task to be segmented, this part of the analysis included exclusively the pressing and slow tapping tasks.

For the pressing task, the four mechanical trigger points were defined as 1) the movement onset –the real-time mechanical onset (force level exceeding 1.3N); 2) the end of the force build-up –the detected end of the force build-up that was defined as the moment when the pressing force exceeded the maximally detectable force of the device (4.4N) or –if the maximally attained force level was below 4.4N- the sign of the slope changed from at least two consecutive positive signs to 0 or negative; 3) the start of releasing –the detected start of releasing that was defined as the moment when the force fell below 4.4 N or –if the maximally attained force level was below 4.4N- the sign of the slope changed from 0 or positive to at least two consecutive negative signs; 4) and the movement offset – the real-time mechanical offset (force level falling below 1.3N).

For the tapping task, the four mechanical trigger points were defined based on the lower and upper photoelectric sensors as 1) movement onset –the time when the index finger had reached the height of the lower photoelectric sensor; 2) the end of finger extension – the time when the index finger had been extended up to the height of the upper photoelectric sensor; 3) the start of finger flexion – the time when the index finger had been lowered below the upper photoelectric sensor; 4) the movement offset –the time when the index finger had been lowered below the lower photoelectric sensor.

##### 1.2 The general delay from mechanical trigger points to trigger points defined at the level of the cortex

Importantly, to estimate the timing of the transition times at the cortex level at the best possible accuracy, we adjusted the trigger points by considering the influence of general mechanical, electromechanical delays and the corticomuscular conduction time across subjects in pressing and tapping events. The electromechanical delay is the time delay between the onset of muscle activation and the measurable movement, reflecting both electrochemical and mechanical processes, previously demonstrated to be around 50ms (Cavanagh and Komi, 1979). And the corticomuscular conduction time was defined as the time lag from the activation of the motor cortex to the activation of muscle, which was measured as 20ms in general, as demonstrated by the TMS experiment (Samii *et al.*, 1998).

For practical purposes, movement onset and offset were defined by the time of passing the lower force threshold for pressing tasks or the activation of the lower light beam sensor for tapping tasks. The mechanical delay was defined as the delay from the actual movement onset/offset to the practically defined mechanical events as determined by the force threshold or photoelectric sensor signal.

We used kinetic signals recorded from the force transducer in both pressing tasks and slow tapping tasks to assess the actual movement onset and offset objectively. For each subject, the digital signals derived from the force transducer were first segmented into 3sec epochs (-1s before the device-detected mechanical onset/offset and 2s after the device-detected mechanical onset/offset). The values representing signals of the four light beam sensors were set to NaN. Then the signals were interpolated to bring the sampling rate to the same value (2kHz). An averaged digital signal across epochs was calculated for each subject. Because the frequency of the movements for pressing and slow tapping tasks was considerably lower than 5Hz for our cohort, the signals were smoothed through a low pass Butterworth filter at 10Hz (2nd order) to avoid interference from high-frequency activities.

We determined the actual movement onset for pressing movements when pressure acceleration reached the maximum and the movement offset as the time when pressure acceleration reached the minimum. To this end, we calculated the second derivative of the digital signals in every 10ms window with a 5ms time step. The movement onset was then determined at the location of the peak (local maximum) close to 0ms (the device-detected mechanical onset), or the location of the valley (local minimum) close to 0ms (the device-detected mechanical offset), as shown in Fig. S1A&B (left panel), respectively. The mechanical delay for each subject on the movement onset/offset is given by the time differences between the actual onset /offset to the 0ms (the real-time detected mechanical onset/offset). The general mechanical delay was then calculated as the averaged mechanical delay across subjects. In this way, the general mechanical delay at the movement onset was 43ms (Fig. S1A, right panel), while the general mechanical delay at movement offset was 9.8ms (Fig. S1B, right panel).

For slow tapping movements, we determined the actual movement onset when the pressure rate reached the minimum, which indicated that the finger had left the pressure plate (One subject was excluded from the analysis because of the loss of the online records of the force transducer). Therefore, we calculated the first derivative of the digital signals in every 10ms window with a 5ms time step. The movement onset was then determined on the valley (local minimum) close to 0ms (the device-detected mechanical onset), as shown in Fig. S1C (left panel). Therefore, the general mechanical delay across subjects at the movement onset was calculated to be 60ms (Figure. S1C, right panel). As for the movement offset, we checked when the finger started to touch the pressure plate. We found a quick (< 10ms) deflection of the force signal right after the finger flexion had crossed the lower light beam sensor (Fig. S1D). We neglected this delay at the movement offset because of its brevity.

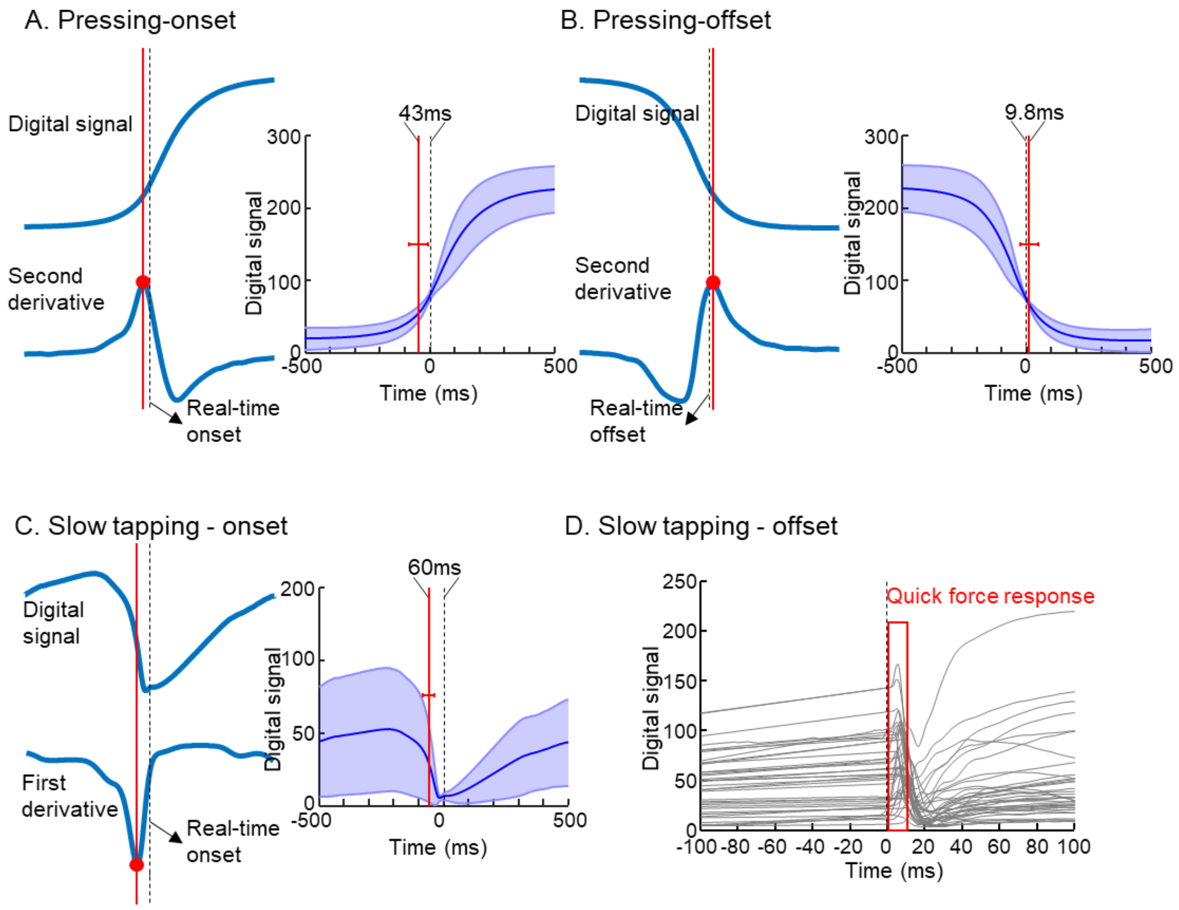

**Figure S1. Estimation of mechanical delay in pressing and slow tapping tasks via digital signals from force transducer.** A. Mechanical delay at the movement onset of pressing task. Left, the adjusted movement onset was determined at the local maximum of the second derivative of the digital signals from the force transducer for each subject. Right, he general mechanical delay was estimated by the time difference between the real-time mechanical onset (0ms point) and the averaged moments for adjusted movement onset across subjects. B. Mechanical delay at the movement offset of pressing task. Left, the adjusted movement onset was determined at the local maximum of the second derivative of the digital signals from the force transducer for each subject. Right, the general mechanical delay was estimated by the time difference between the real-time mechanical offset (0ms point) and the averaged movements for adjusted movement offset across subjects. C. Mechanical delay at the movement onset of slow tapping task. Left, the adjusted movement onset was determined at the local minimum of the first derivative of the digital signals from the force transducer for each subject. Right, the general mechanical delay was estimated by the time difference between the real-time mechanical onset (0ms point) and the averaged moments for adjusted movement onset across subjects. D. Mechanical delay at the movement offset of the slow tapping task. The line plot showed interpolated digital signals of all subjects. Most of the subjects showed a quick and large force response right after the finger flexion interrupted the lower light sensor (real-time mechanical offset, 0ms point). The period from real-time mechanical offset to the deflection of the force transducer was neglected because of its brevity.

##### 1.3 The definition of adjusted trigger points at the level of the cortex

Overall, in the pressing task, the general delay in total from the adjusted movement onset at the cortical level to the real-time mechanical onset of pressing was estimated as 113ms (20ms + 50ms + 43ms). The delay from the adjusted movement offset to the real-time mechanical offset of pressing was estimated as 60ms (20ms + 50ms -10ms). Besides, due to the force saturation (the maximum force that can be detected is 4.5N) during the pressing in most cases, the realistic ending of pressing and start of releasing cannot be acquired, the delays for the two trigger points between the movement onset and offset only included the general electromechanical delay and the corticomuscular conduction time, which are 70ms. Therefore, for the pressing task, also considering the 50ms time resolution of the movement-PAC values, the 4 trigger points at the cortex level were defined as: 1) the movement onset – 150ms before the real-time mechanical onset (force level exceeding 1.3N); 2) the end of the pressing build up – the closest time point to the 70ms before the detected end of the force build-up (the moment when the pressing force exceeded the maximally detectable force of the device (4.4N) or –if the maximally attained force level was below 4.4N- the sign of the slope changed from at least 2 consecutive positive signs to 0 or negative); 3) the start of releasing – the closest time point to 70 ms before the detected start of releasing (the moment when the force fell below 4.4N or – if the maximally attained force level was below 4.4N - the sign of the slope changed from 0 or positive to at least 2 consecutive negative signs); 4) and the movement offset – the closest time point to 60ms before the mechanical offset (force level falling below 1.3N).

In the slow tapping task, the delay in total from the adjusted movement onset at the cortical level to real-time mechanical onset of tapping was estimated as 130ms (20ms + 50ms + 60ms). The delay from the adjusted movement offset to the device-detected offset of tapping only contains the general electromechanical delay and the corticomuscular conduction time, which in total was 70ms. Besides, because the device did not record the whole trace of the finger tapping when the finger left the pressing board, the delays for the two trigger points between the movement onset and offset only included the general electromechanical delay and the corticomuscular conduction time, which are 70ms. Therefore, for the slow tapping task, also considering the time resolution (50ms) of movement-related PAC time series, the 4 trigger points at the cortex level were defined based on the lower and upper photoelectric sensors as: 1) movement onset – 150ms before the time when the index finger had been extended higher than the lower photoelectric sensor. 2) the end of finger extension – the time point closest to 70ms before the time when the index finger had been extended higher than the upper photoelectric sensor. 3) the start of finger flexion – the time point closest to 70ms before the time when the index finger had been lowered below the upper photoelectric sensor. 4) the movement offset – the time point closest to 70ms before the time when the index finger had been lowered below the lower photoelectric sensor.

### Supplementary tables

#### Supplementary Table 1. Characteristics of patients with Parkinson's disease

| ID | Sex | Disease Duration, years | Clinically more affected body side/ MDS-UPDRS III hemi-body-scores | Total MDS-UPDRS III (medication off) | L-Dopa equivalent dose mg/day |
|---|---|---|---|---|---|
| **01** | Male | 4 | Right/7 | 15 | 310 |
| **02** | Female | 1 | Left/5 | 6 | 210 |
| **03** | Male | 1 | Left/10 | 22 | 400 |
| **04** | Female | 3 | Left/6 | 10 | 152 |
| **05** | Male | 1 | Left/8 | 12 | 500 |
| **06** | Male | 4 | Right/10 | 30 | 735 |
| **07** | Male | 12 | Right/12 | 20 | 682.5 |
| **08** | Female | 11 | Left/11 | 24 | 400 |
| **09** | Male | 3 | Left/10 | 30 | 525 |
| **10** | Male | 2 | Right/5 | 12 | 100 |
| **11** | Female | 4 | Left/13 | 32 | 300 |
| **12** | Female | 6 | Left/7 | 19 | 955 |
| **13** | Male | 12 | Left/11 | 35 | 1395 |
| **14** | Male | 2 | Right/6 | 11 | 355 |
| **15** | Female | 3 | Left/15 | 26 | 930 |
| **16** | Male | 6 | Right/10 | 20 | 400 |
| **17** | Male | 17 | Right/12 | 29 | 1185 |
| **18** | Male | 2 | Right/13 | 21 | 520 |
| **19** | Male | 6 | Right/8 | 14 | 930 |

the table is adapted from Gong et al., Brain 2021, with permission (Gong *et al.*, 2021)

#### Supplementary Table 2. Number of epochs in each condition

| | Resting | Pressing | Slow tapping, FB- | Slow tapping, FB+ | Fast tapping, FB- | Fast tapping, FB+ | Sign-rank p-value |
|---|---|---|---|---|---|---|---|
| **Patients** | 105.6±12.1 | 104.9±26.3 | 110.2±26.3 | 110.8±25.5 | 101.4±15.2 | 99.4±14.8 | >0.2 |
| **Controls** | 108.5±5.3 | 103.6±28.6 | 101.1±30.6 | 103.2±31.4 | 103.3±15.7 | 100.6±18.2 | >0.06 |
| **Rank-sum p-value** | 0.36 | 0.99 | 0.21 | 0.39 | 0.34 | 0.35 | |

FB-: without feedback; FB+: with feedback

#### Supplementary Table 3. Number of ICA components derived from ROI source signals

| | | Resting | Pressing | Slow tapping, FB- | Slow tapping, FB+ | Fast tapping, FB- | Fast tapping, FB+ |
|---|---|---|---|---|---|---|---|
| **PMC** | **Patients** | 10.5±0.6 | 10.2±0.7 | 10.2±0.9 | 10.2±0.9 | 10.0±0.9 | 10.1±0.9 |
| | **Controls** | 10.7±0.7 | 10.5±0.5 | 10.8±0.8 | 10.6±0.7 | 10.4±0.8 | 10.3±0.9 |
| **M1** | **Patients** | 10.5±0.5 | 10.5±0.5 | 10.4±0.8 | 10.5±0.8 | 10.4±0.8 | 10.5±0.9 |
| | **Controls** | 10.7±0.7 | 10.6±0.8 | 10.5±0.8 | 10.5±0.8 | 10.3±0.9 | 10.3±0.9 |
| **BA3** | **Patients** | 10.6±0.7 | 10.7±0.7 | 10.6±1.0 | 10.5±0.9 | 10.4±1.0 | 10.5±0.9 |
| | **Controls** | 10.9±0.6 | 10.8±0.6 | 10.8±0.7 | 10.7±0.8 | 10.7±0.7 | 10.6±0.7 |
| **BA1&2** | **Patients** | 10.8±0.7 | 10.6±0.8 | 10.7±0.9 | 10.7±0.9 | 10.6±0.9 | 10.7±0.7 |
| | **Controls** | 11.0±0.7 | 10.9±0.7 | 10.9±0.8 | 10.9±0.7 | 10.7±0.8 | 10.8±0.8 |

PMC: Premotor cortex; M1: Primary motor cortex; BA3: Primary somatosensory cortex; BA1&2: Primary somatosensory complex; FB-: without feedback; FB+: with feedback
